# SARS-CoV-2 Innate Effector Associations and Viral Load in Early Nasopharyngeal Infection

**DOI:** 10.1101/2020.10.30.20223545

**Authors:** Theodore G Liou, Frederick R Adler, Barbara C Cahill, David R Cox, James E Cox, Garett J Grant, Kimberly E Hanson, Stephen C Hartsell, Nathan D Hatton, My N Helms, Judy L Jensen, Christiana Kartsonaki, Yanping Li, Daniel T Leung, James E Marvin, Elizabeth A Middleton, Sandra M Osburn-Staker, Kristyn A Packer, Salika M Shakir, Anne B Sturrock, Keith D Tardif, Kristi Jo Warren, Lindsey J Waddoups, Lisa J Weaver, Elizabeth Zimmerman, Robert Paine

**Affiliations:** Division of Respiratory, Critical Care and Occupational Pulmonary Medicine, Department of Internal Medicine, School of Medicine, University of Utah, Salt Lake City, UT, USA; Center for Quantitative Biology, University of Utah, Salt Lake City, UT, USA; Department of Mathematics, College of Science and Department of Biology, College of Biological Sciences, University of Utah, Salt Lake City, UT, USA; Nuffield College, Oxford, UK; Department of Biochemistry, School of Medicine, University of Utah, Salt Lake City, UT, USA; Metabolomics, Proteomics and Mass Spectrometry Core, School of Medicine, University of Utah, Salt Lake City, UT, USA; Division of Infectious Diseases, Department of Internal Medicine, School of Medicine, University of Utah, Salt Lake City, UT, USA; Department of Pathology, University of Utah, Salt Lake City, UT, USA; ARUP Laboratories, Salt Lake City, UT, USA; Division of Emergency Medicine, Department of Surgery, University of Utah, Salt Lake City, UT, USA; Clinical Trial Service Unit & Epidemiological Studies Unit and Medical Research Council Population Health Research Unit, Nuffield Department of Population Health, University of Oxford, Oxford, UK; Flow Cytometry Core Laboratory, University of Utah Health, Salt Lake City, UT, USA; Department of Veterans Affairs Medical Center, Salt Lake City, UT, USA

**Author notes:** Correspondence should be addressed to TGL.

## Abstract

To examine innate immune responses in early SARS-CoV-2 infection that may change clinical outcomes, we compared nasopharyngeal swab data from 20 virus-positive and 20 virus-negative individuals. Multiple innate immune-related and *ACE-2* transcripts increased with infection and were strongly associated with increasing viral load. We found widespread discrepancies between transcription and translation. Interferon proteins were unchanged or decreased in infected samples suggesting virally-induced shut-off of host anti-viral protein responses. However, IP-10 and several interferon-stimulated gene proteins increased with viral load. Older age was associated with modifications of some effects. Our findings may characterize the disrupted immune landscape of early disease.

---

The coronavirus disease 2019 (COVID-19) pandemic due to severe acute respiratory syndrome (SARS) coronavirus-(CoV)-2 infection has afflicted millions following first reports of a novel viral pneumonia on 31 December 2019 in Wuhan, China.^1^ Rapid publication of molecular characterization of the virus^2–4^ enabled multiple instances of successful detection, tracking and containment using public health measures.^5–10^ In most localities, however, viral spread has been rapid, widespread and devastating, facilitated by high rates of transmission often before the appearance of symptoms and followed by elevated rates of morbidity and mortality.

Silent infection with rapid viral replication allows asymptomatic person-to-person infection.^5–7,11^ Nasopharyngeal viral loads peak when upper airway symptoms appear,^5,12^ and the size of the initial viral innoculum may determine the rapidity of onset and severity of the subsequent clinical syndrome.^13^ Over days, sometimes weeks, the infection may extend locally to involve the lower respiratory tract. For a substantial portion of infected patients, initial fever, dry cough, myalgias and anosmia progress to dyspnea, hypoxemic respiratory failure and the acute respiratory distress syndrome (ARDS).^14–17^

Pathophysiological disturbances of infection are multi-faceted and can be severe. Some of the laboratory findings include neutrophilia, lymphopenia, decreased T helper and suppressor cells, abnormal platelet function and coagulation cascade derangements. Substantial numbers of patients progress to clinically severe vasculitides, thromboembolic disorders, pneumonia or ARDS.^18,19^

In patients who develop ARDS, dysfunctional innate immune and systemic inflammatory responses include marked elevations in systemic inflammatory cytokines such as interleukin-(IL)-6 and tumor necrosis factor-(TNF)-α in both serum and airway secretions as well as seemingly insufficient anti-viral interferon (IFN) responses, all of which have been described as a cytokine storm.^20–24^ Disruptions in human biochemical marker responses begin early in disease, as recent evaluations of immunoglobulins and other markers of immune responses in serum demonstrate.^25^ Nevertheless, much remains unexplored, particularly in the airway compartment, the initial site of infection.

Part of the challenge to developing a treatment or prevention response is to elucidate the range of innate immune responses to SARS-CoV-2 infection that may precede the cytokine storm seen in severe disease.^24,26^ Evaluations in ARDS, especially due to severe respiratory viral infections such as Influenza A,^23,27,28^ Middle Eastern Respiratory Syndrome,^29–31^ SARS-CoV^32–37^ and other highly pathogenic coronaviruses such as Human CoV-(HCoV)-Erasmus Medical Center (HcoV-EMC)^38–40^ highlight the broad collection of biomarkers that may potentially be useful in SARS-CoV-2 infection, especially prior to fulminant disease, uncontrolled systemic inflammation and ARDS.

Coronaviruses that cause severe human disease are remarkable for their ability to evade innate immune defenses and to promote dysfunctional responses that appear before cytokine storm.^41,42^ For example, IFN responses are critically important for anti-viral defense,^43^ yet there is no detectable native human IFN response to SARS-CoV.^38^ No fewer than 11 of 28 known SARS-CoV proteins interfere with signaling cascades that produce IFN proteins after endosomal or cytoplasmic viral detection.^44^ *In vitro* studies of various human cell types show that SARS-CoV efficiently suppresses transcription of IFN genes but selectively allows expression and translation of other genes.^45–47^ Bronchoalveolar lavage studies of epithelial cells obtained from severely ill patients following SARS-CoV-2 infection suggest a pathophysiology more consistent with HCoV-EMC or MERS-CoV than SARS-CoV.^21^ However both SARS-CoV and SARS-CoV-2 stimulate inflammatory signals via nuclear factor κB (NFκB)^44^ that recruit polymorphonuclear neutrophils and other immune effector cells to the lung, releasing proteases that may dramatically further increase viral cell entry.^48–50^ IFN-α and IFN-β treatments that bypass some evasion strategies^21^ have been proposed to counter both SARS-CoV viruses.^51–53^ However, we lack efficacy and safety trials free of observer bias,^54^ and no published human data exists for IFN-λ therapy.

To supplement the growing information on responses early in infection, we undertook an observational study of deidentified nasopharyngeal swab samples from patients presenting at drive-through testing centers for evaluation of symptoms potentially due to SARS-CoV-2 infection. We selected proteins involved in different steps of human cellular responses to viral invasion for quantitative measurements by multiple methods to understand the impact of targeting by viral evasion activities.^29,42,44^ We selected and measured factors important for understanding viral entry, intracellular detection of viral invasion, production of pro-inflammatory signals, systemic inflammatory agents and multiple IFN and IFN-stimulated gene (ISG) responses relative to viral loads to better understand the immune landscape of patients with early disease.

## Results

### Study Population

We evaluated 40 samples from individuals, evenly divided into 20 positive and 20 negative detection results for SARS-CoV-2. Samples were deidentified but annotated by age (median 46.5 years, range 11-90) and sex (17 females, 42.5%). Older patients were more likely to be male and negative for SARS-CoV-2 detection (Supplementary Figure 1 and Supplementary Table 1a and 1b). This limited demographic information suggested that further evaluation of statistical relationships in our sample set required testing adjustments for age, sex or both to avoid confounding.

Samples included in our study were randomly selected from those collected from April-June of 2020 from patients who may have come from nine states within the Mountain West of the United States. During this period, positive results were reported for about 9-10% of tested patients. Among the positive test patients, about 10% eventually required hospitalization for COVID-19 with less than 50% of those hospitalized suffering respiratory failure, ARDS or succumbing to severe disease. While we know this context for our samples, the specific outcomes for individual patients in our study are unknown.

### Viral Load

Using RT-PCR results, we estimated fold-change in mRNA expression of SARS-CoV-2 small envelope protein E1 (Odds Ratio [OR] = 10.8 × 10^6^, 95% Confidence Interval [CI] = 8.37 × 10^5^-1.40 × 10^8^, *p* < 0.001) and nucleocapsid protein N1 (OR = 5.1 × 10^7^, CI = 4.5 × 10^6^-5.9 × 10^8^, *p* < 0.001) relative to expression in patients without infection. We selected primers^55^ for *E1* originally from Charité, Germany^4^ and *N1* from the US CDC.^56^ Both *E1* and *N1* mRNA fold changes gave virtually total discrimination between patients with and without infection diagnosed by clinical testing for SARS-CoV-2 infection using qualitative RT-PCR.

### Viral Entry

We measured two human protein transcripts important for understanding SARS-CoV-2 cell entry, *angiotensin converting enzyme-2* (*ACE-2*), which is essential for entry of SARS-CoV-2 and SARS-CoV,^48,57^ and *transmembrane protease, serine-2* (*TMPRSS-2*) which enhances cell entry up to a thousand-fold.^48,49^ *ACE-2* mRNA was increased three-fold in patients with infection, and the fold-change results were strongly associated with viral load. *TMPRSS-2* mRNA expression, however, was not associated with infection nor viral load (Figure 1).

**Figure 1.**
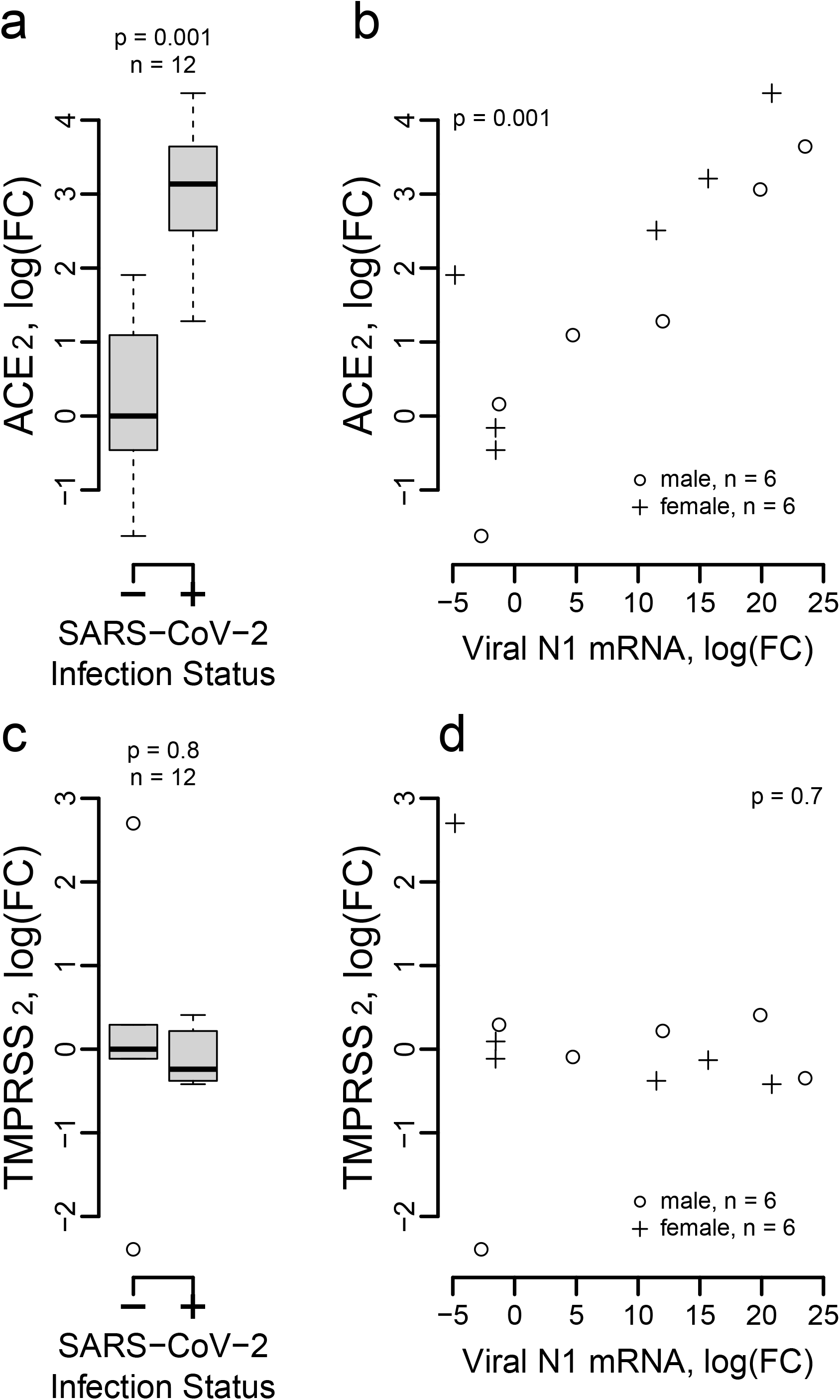
Association of *ACE2* but not *TMPRSS2* Expression with SARS-CoV-2 Infection. **a**. *ACE-2* mRNA is increased approximately three-fold in patients with SARS-CoV-2 infection over *ACE-2* mRNA expression in patients without infection, and **b**. the increase in expression is associated with viral load (OR = 1.16, CI = 1.1-1.23, *p* < 0.001). However, the expression of *TMPRSS-2* is **c**. neither increased nor decreased with infection and **d**. is not associated with viral load. Adjustments for age and sex were not significant for either molecule. In each panel, **a** and **c**, there are six infected and six non-infected status patients.

### Viral Detection Signaling

We examined transcription signals for two genes in the signaling pathway downstream of viral detection important for IFN responses, *TNF-associated factor-binding kinase-1 associated with inhibitor of NFκB* (*TBK-1*) and *Stimulator of IFN genes-*(*STING*)-*1* for six patients with positive detection of SARS-CoV-2 and six patients with negative detection. The mRNA expressions of *TBK-1* and *STING-1* were not associated with infection (Supplementary Figure 2).

### Inflammatory Responses to SARS-CoV-2 Infection

We found increased mRNA expression of *IL-8, IFN-γ-induced protein-*(*IP*)-*10* and *TNF-α* in SARS-CoV-2-infected individuals (Figure 2a-c). Moreover, we found that there was a strong association between viral load and the level of mRNA expression of these innate immune effector moleculesI agonized (Figure 2f-h).

**Figure 2.**
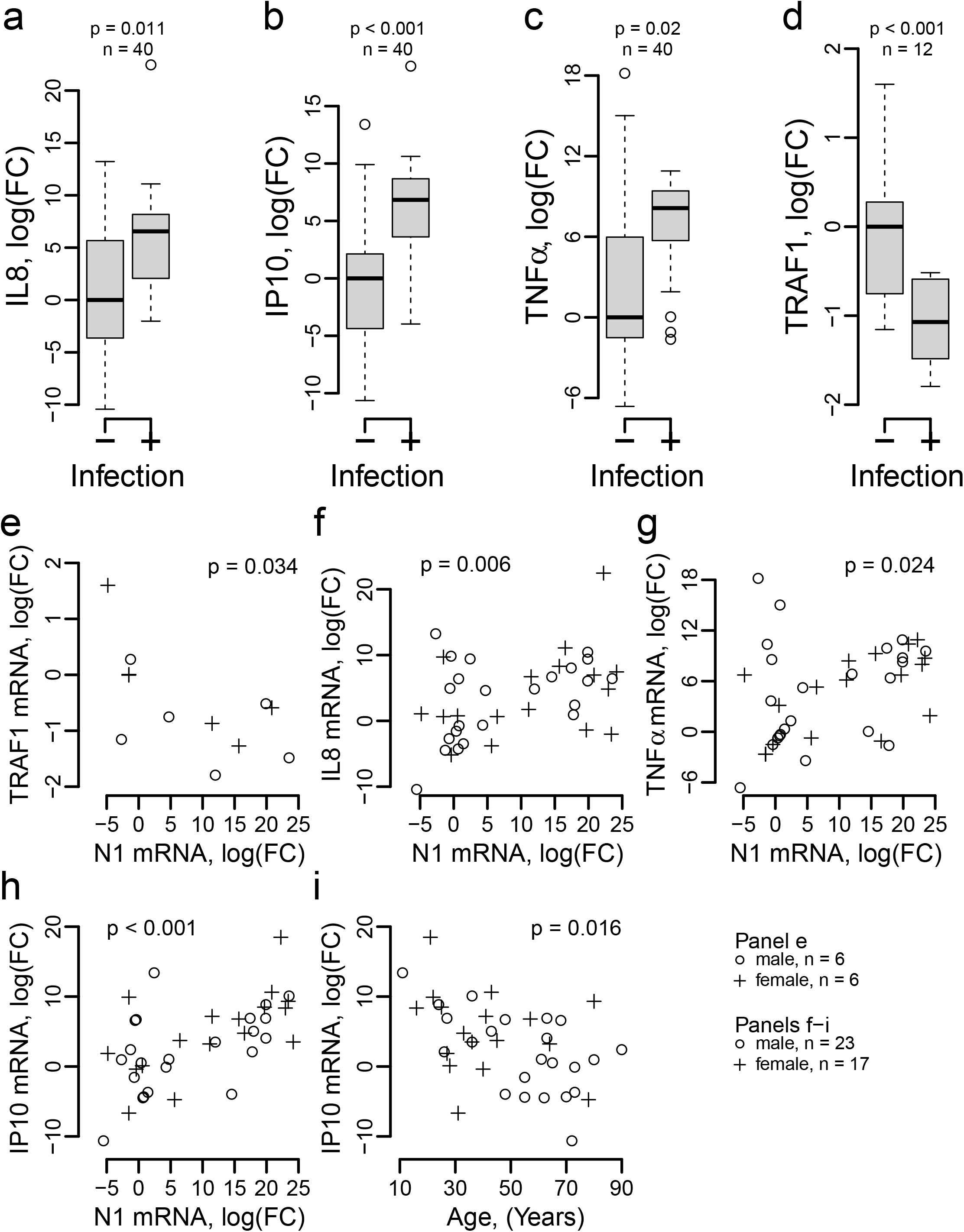
Transcripts for Genes Associated with Inflammation. Transcription of **a**. *IL-8*, **b**. *IP-10*, **c**. *TNF-α* mRNA are all increase approximately 6-fold while transcription of **d**. *TRAF-1* is reduced. When assessed for relationship with viral load, decreasing **e**. *TRAF-1* mRNA is associated with increasing viral *N1* protein mRNA while increasing mRNA for **f**. *IL-8*, **g**. *TNF-α* (less strongly) and **h**. *IP-10* are associated with increasing viral *N1* protein mRNA. In contrast, decreasing **i**. *IP-10* mRNA is associated with increasing age. Similar relationships are seen with viral *E1* protein transcripts (not shown). In each panel, **a**-**c**, there are 20 infected and 20 non-infected status patients. In panel **d**, there are 6 infected and 6 non-infected.

Viral entry, detection and signaling may lead to a systemic inflammatory response via *NFκB* activity which may potentially be augmented by *TNF receptor-associated factor-*(*TRAF*)*-1* activity.^58,59^ We found a large reduction in *TRAF-1* mRNA (Figure 2d). The reduction in *TRAF-1* was inversely associated with expression of both viral protein *E1* mRNA (OR = 0.945, CI = 0.906-0.986, *p* = 0.025) and *N1* mRNA (OR = 0.947, CI = 0.907-0.989, *p* = 0.034, Figure 2e). We found *NFκB-1* and *NFκB-2* mRNA transcripts were not significantly changed compared to uninfected status (Table 1a). Other downstream immune effectors, *granulocyte-macrophage colony stimulating factor* (*GM-CSF*), *IL-6* and *IL-10* mRNA were not increased (Table 1a).

**Table 1a.**
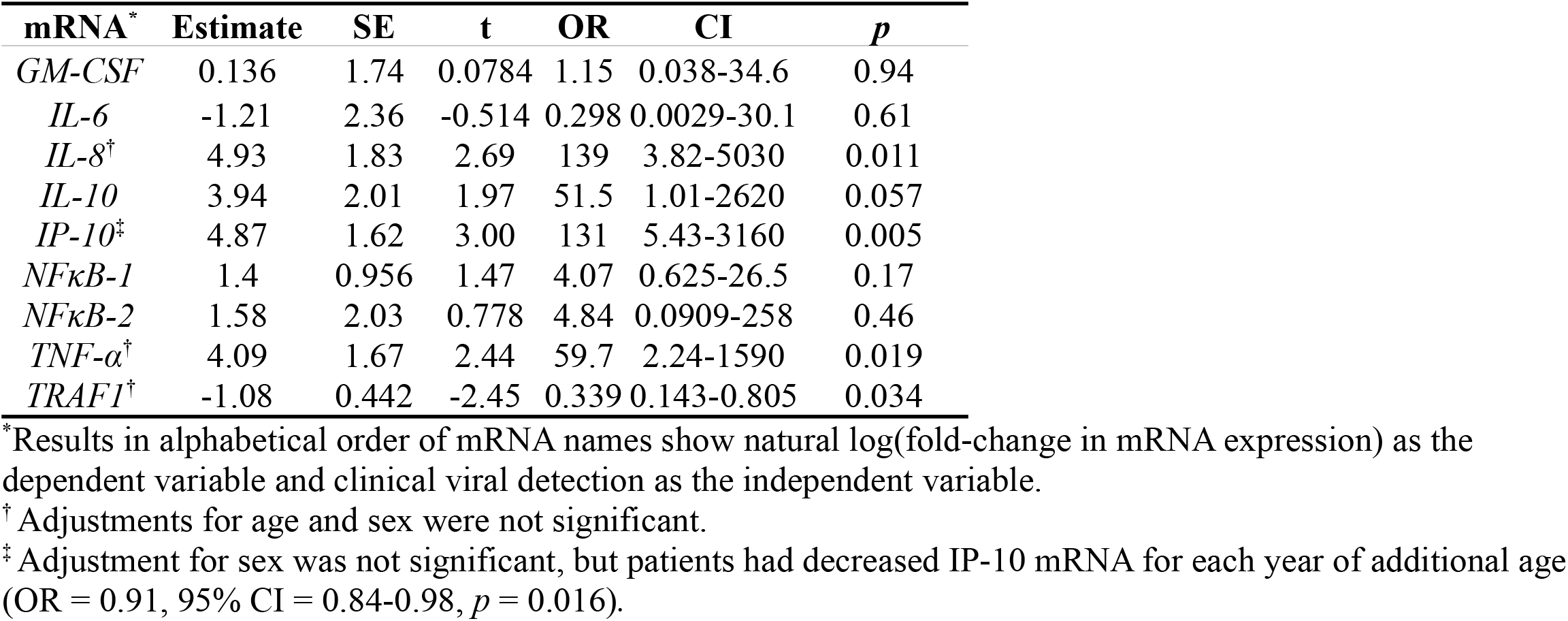
Systemic Inflammatory mRNA Response to SARS-CoV-2 Infection.

| mRNA<sup>*</sup> | Estimate | SE | t | OR | CI | p |
| --- | --- | --- | --- | --- | --- | --- |
| *GM-CSF* | 0.136 | 1.74 | 0.0784 | 1.15 | 0.038-34.6 | 0.94 |
| *IL-6* | -1.21 | 2.36 | -0.514 | 0.298 | 0.0029-30.1 | 0.61 |
| *IL-8*<sup>†</sup> | 4.93 | 1.83 | 2.69 | 139 | 3.82-5030 | 0.011 |
| *IL-10* | 3.94 | 2.01 | 1.97 | 51.5 | 1.01-2620 | 0.057 |
| *IP-10*<sup>‡</sup> | 4.87 | 1.62 | 3.00 | 131 | 5.43-3160 | 0.005 |
| *NFκB-1* | 1.4 | 0.956 | 1.47 | 4.07 | 0.625-26.5 | 0.17 |
| *NFκB-2* | 1.58 | 2.03 | 0.778 | 4.84 | 0.0909-258 | 0.46 |
| *TNF-α*<sup>†</sup> | 4.09 | 1.67 | 2.44 | 59.7 | 2.24-1590 | 0.019 |
| *TRAF1*<sup>†</sup> | -1.08 | 0.442 | -2.45 | 0.339 | 0.143-0.805 | 0.034 |

<sup>\*</sup>Results in alphabetical order of mRNA names show natural log(fold-change in mRNA expression) as the dependent variable and clinical viral detection as the independent variable.
<sup>†</sup>Adjustments for age and sex were not significant.
<sup>‡</sup>Adjustment for sex was not significant, but patients had decreased IP-10 mRNA for each year of additional age (OR = 0.91, 95% CI = 0.84-0.98, *p* = 0.016).

Protein measurements using bead-based multiplex immunoassays (BioLegend) for systemic inflammatory markers matching many of the mRNAs measured (plus IL-1b and IL-12p70) revealed no significant changes with infection and no significant associations with viral load with one exception. IP-10 protein was increased nearly four fold above measured control values (Table 1b), and the log of concentration was strongly associated with viral load (OR = 1.09 per unit of log unit of viral *N1* mRNA fold-change, CI = 1.06-1.12, *p* < 0.001).

**Table 1b.** Protein production of inflammatory markers (log[pg/ml]) with SARS-CoV-2 Infection.

| Protein<sup>*,†</sup> | Estimate | SE | t | OR | CI | p |
| --- | --- | --- | --- | --- | --- | --- |
| GM-CSF | -0.00644 | 0.0060 | -1.07 | 0.994 | 0.982-1.01 | 0.29 |
| IL-1b | -0.0594 | 0.0358 | -1.66 | 0.942 | 0.878-1.01 | 0.10 |
| IL-6 | 0.116 | 0.113 | 1.02 | 1.12 | 0.899-1.4 | 0.32 |
| IL-8 | 0.0976 | 0.457 | 0.214 | 1.1 | 0.45-2.7 | 0.83 |
| IL-10 | 0.106 | 0.0846 | 1.25 | 1.11 | 0.942-1.31 | 0.22 |
| IL-12p70 | -0.0888 | 0.078 | -1.14 | 0.915 | 0.785-1.07 | 0.26 |
| IP-10 | 1.32 | 0.302 | 4.36 | 3.74 | 2.07-6.77 | <0.001 |
| TNF-α | 0.0265 | 0.0265 | 1.00 | 1.03 | 0.975-1.08 | 0.32 |

<sup>\*</sup>Results show natural log(pg/ml) of each protein as a function of viral detection.
<sup>†</sup>Adjustments for age and sex were not significant.

### IFN Responses to Infection

We found five to six-fold increases in expression of both *IFN-λ1* and *IFN-λ2* mRNA among patients with detection of SARS-CoV-2 (n=20) compared to patients without detection of virus (n=20) (Table 2a, Figure 3a and 3c). There were no other significant increases in *IFN* mRNA. The increases in *IFN-λ1* and *IFN-λ2* mRNA production were strongly associated with viral load (Figure 3e and 3f).

**Table 2a.** IFN Response to SARS-CoV-2 Infection.

| mRNA* | Estimate† | SE | t | OR | CI | p |
| --- | --- | --- | --- | --- | --- | --- |
| *IFN-α2* | 1.17 | 0.993 | 1.17 | 3.21 | 0.458-22.5 | 0.25 |
| *IFN-β1* | 0.88 | 1.16 | 0.756 | 2.41 | 0.246-23.6 | 0.45 |
| *IFN-γ* | 1.89 | 1.03 | 1.84 | 6.65 | 0.883-50.1 | 0.074 |
| *IFN-λ1* | 4.26 | 1.18 | 3.62 | 71 | 7.07-713 | <0.001 |
| *IFN-λ2* | 3.69 | 1.2 | 3.09 | 40.2 | 3.86-419 | 0.004 |
| *IFN-λ3* | -0.991 | 2.67 | -0.371 | 0.371 | 0.00196-70.2 | 0.71 |

\*Results are show natural log(fold-change in mRNA expression) as a function of viral detection.
†Adjustments for age and sex were not significant for any model.

**Figure 3.**
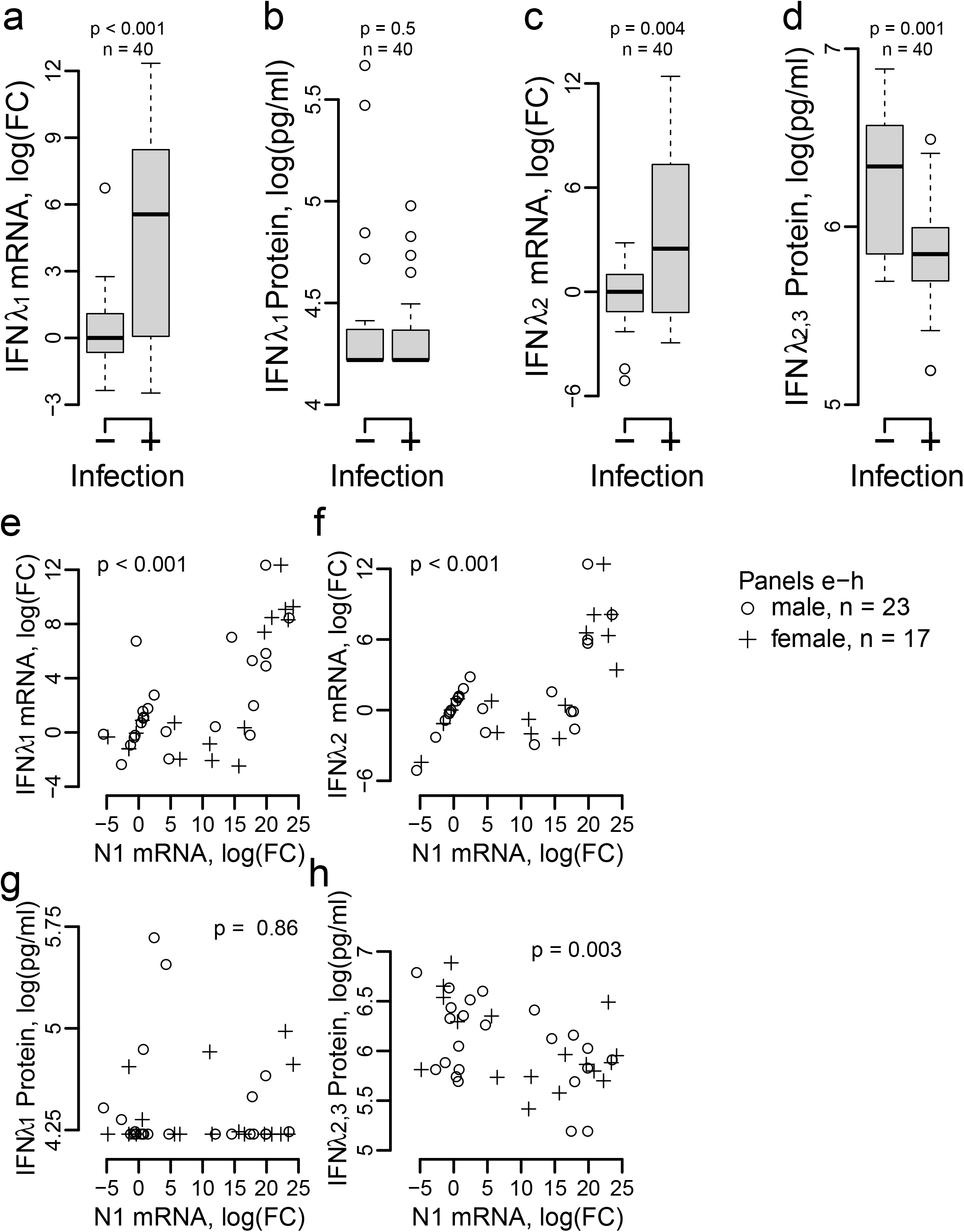
Transcripts for Genes Associated with Interferons. Transcription of **a**. *IFN-λ1* is increased nearly 6-fold although **b**. IFN-λ1 protein is unchanged with infection. Transcription of **c**. *IFN-λ2* is increased about 3-fold but **d**. IFN-λ2 protein is decreased even if the combined measurement of IFN-λ2 and IFN-λ3 proteins are attributed solely to IFN-λ2. The mRNA fold changes for **e**. *IFN-λ1* and **f**. *IFN-λ2* are directly associated with increasing viral *N1* protein mRNA. For **g**. IFN-λ1 protein there is no association with viral *N1* protein mRNA, but for **h**. combined IFN-λ2 and IFN-λ3 protein measurement, there is an inverse relationship with viral *N1* protein mRNA. For **e**.-**h**., substitution of viral *E1* mRNA produced similar relationships and figures. In each panel, **a**-**d**, there are 20 infected and 20 non-infected status patients.

Despite increased mRNA expression for some of the IFNs, protein measurements showed reductions in IFN-α2, IFN-γ and IFN-λ2,3 in patients with viral infection that averaged 66%, 49% and 40%, respectively, relative to control patients (Table 2c).

### ISG Responses to Infection

We prospectively selected four ISGs to evaluate because of their importance in defense against RNA viruses^60^: GTP-binding Myxovirus protein (*MX-1*),^61–65^ IFN-induced proteins with tetratricopeptide repeats (*IFIT*),^66^ IFN-induced transmembrane protein (*IFITM*)^66,67^ and Tetherin (*BST-2*).^68,69^ *MX-1* and *Tetherin* mRNA were both increased in infected patients with moderate statistical significance while two *IFIT* mRNAs were greatly and significantly increased (Table 2d). Focused proteomic examination of proteins extracted from samples using data independent acquisition (DIA) mass spectrometry detected an association between positive clinical testing for SARS-CoV-2 and IFIT-1, IFIT-3 and Tetherin proteins with a borderline finding for MX-1 protein (Table 2e). Proteomic examination using data dependent acquisition (DDA) mass spectrometry, which has less sensitivity but better specificity and precision, detected only a large increase in IFIT-3 protein that was associated with clinical infection detection and increasing viral *N1* mRNA (Figure 4). Transcript and proteome results are based on the same six positive and six negative samples for which we had sufficient mRNA remaining after other studies.

**Figure 4.**
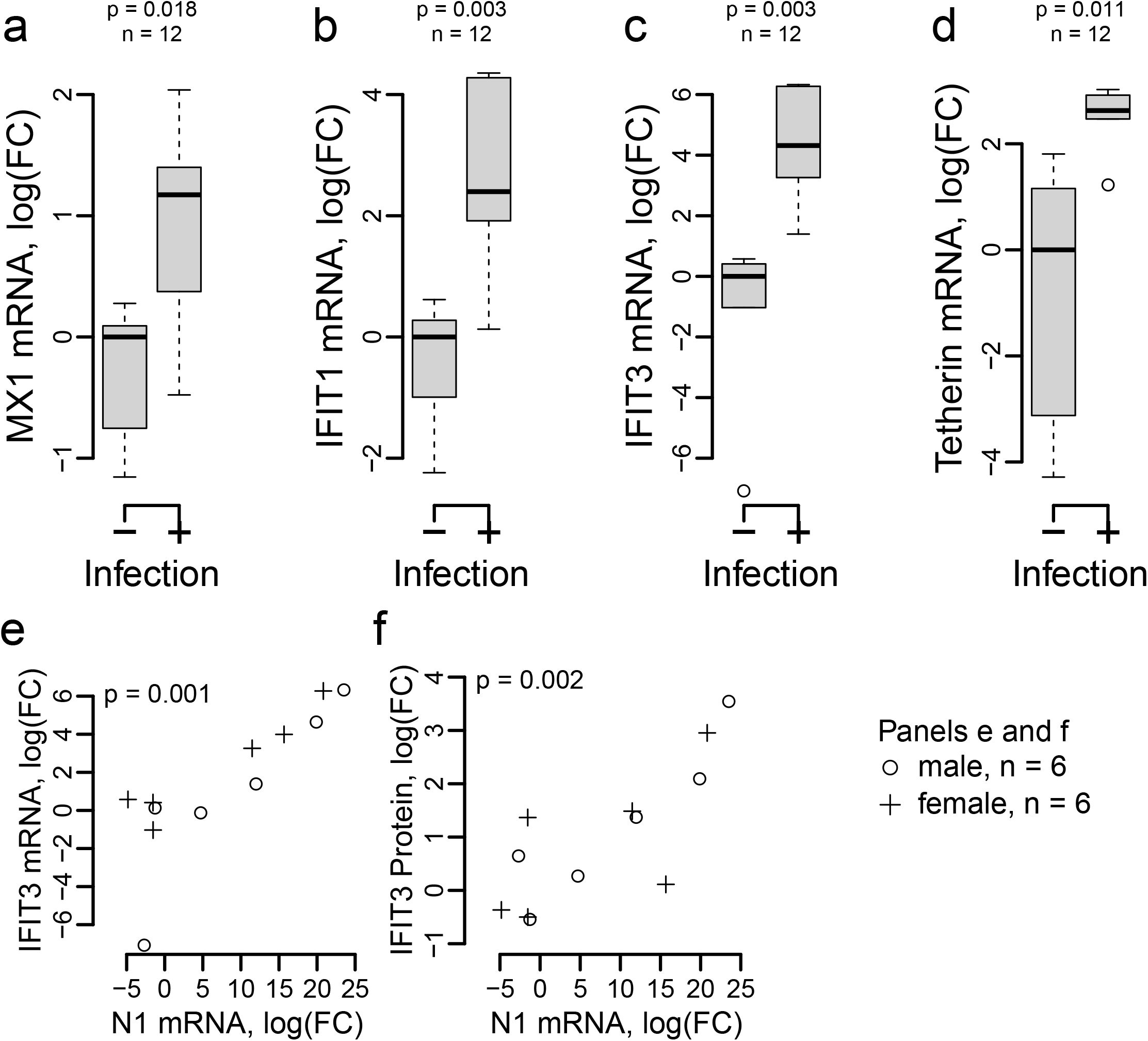
Transcripts for Pre-selected ISGs. Infection with SARS-CoV-2 is associated with increased **a**. *MX-1*, **b**. *IFIT-1*, **c**. *IFIT-3* and **d**. *Tetherin* (*BST-2*) mRNAs. There were significant associations between increasing **e**. *IFIT-3* mRNA and **f**. IFIT-3 protein fold changes and increasing viral *N1* protein mRNA. Protein fold-change for IFIT-3 were measured using DDA mass spectrometry. Similar relationships were seen using viral *E1* protein mRNA as in **e**. and **f**. In each panel, **a**-**d**, there are six infected and six non-infected status patients.

### Model Adjustments with Older Age

The increase in *IP-10* mRNA with infection was lower in older individuals (Figure 2i). All other things being equal, 10 additional years in age were associated with an approximately overall 70% reduction in *IP-10* mRNA compared to controls while 25 additional years in age were associated with an overall 90% reduction, producing a remarkable counter effect to infection itself.

We found a borderline significant effect for IFN-λ2,3. Patients with older ages have slightly lower IFN-λ2,3 per additional year of age with SARS-CoV-2 infection. This small per year effect was associated with a 10% lower IFN-λ2,3 on average for every additional 10 years of age and about 23% lower IFN-λ2,3 for an additional 25 years of age in addition to the approximately 40% reduction associated with SARS-CoV-2 infection (Table 2c).

### Correlations between IFN, Inflammatory and ISG Measurements

We found no strong and significant correlations between any protein and its transcript suggesting widespread translation blockade or rapid protein degradation with one exception. Both *IP-10* mRNA and protein were directly associated with viral load (*N1* protein mRNA), and the correlation between mRNA and protein was moderate with strong significance, *p <* 0.001 (Supplementary Table 2b). In contrast, we found strong correlations within IFN and inflammatory proteins (Supplementary Table 2a) and within IFN gene, inflammatory signaling gene and ISG transcripts (Supplementary Table 2c). Correlations within IFN types, for example, among IFN-λ sub types were exceptionally strong which corresponds to the biology of the IFNs.

### Sensitivity Testing

We performed sensitivity testing for all significant associations between IFN, inflammatory and ISG measurements with viral load (*N1* protein mRNA) by re-examining the relationships after exclusion of patients without infection by SARS-CoV-2. In every case, we found similar relationships between each biomarker and viral load, increasing the confidence in our findings.

Because of the high degree of correlation between some biomarkers (Supplementary Table 2), for example between *IFN-β1* and *IFNα2* transcripts (Spearman correlation coefficient = 1.00, *p* < 0.001), we examined the effect of adding a second biomarker as an adjustment to the relationship between each significant biomarker (*p* < 0.05) with the clinical diagnosis of infection (Tables 1a, 1b, 2a, 2c and 2d) or with viral load (Tables 2b and individually reported results for *ACE2* mRNA, *IP-10* mRNA and IP-10 protein). In every case, we found similar results for the association between the biomarkers reported and infection status or viral load. A number of the biomarker measurements tested as adjustment variables appeared to have independent significant effects suggesting that significant and independent multivariable associations exist, however, our study is too small to report those results with confidence.

**Table 2b.** IFN response associations with viral load (N1 protein).

| mRNA* | Estimate† | SE | t | OR | CI | p |
| --- | --- | --- | --- | --- | --- | --- |
| *IFN-α2* | 0.102 | 0.0495 | 2.06 | 1.11 | 1.01-1.22 | 0.046 |
| *IFN-β1* | 0.0904 | 0.0588 | 1.54 | 1.09 | 0.975-1.23 | 0.13 |
| *IFN-γ* | 0.14 | 0.0507 | 2.75 | 1.15 | 1.04-1.27 | 0.009 |
| *IFN-λ1* | 0.302 | 0.0507 | 5.96 | 1.35 | 1.23-1.49 | <0.001 |
| *IFN-λ2* | 0.285 | 0.0515 | 5.53 | 1.33 | 1.2-1.47 | <0.001 |
| *IFN-λ3* | 0.0531 | 0.138 | 0.384 | 1.05 | 0.804-1.38 | 0.70 |

\*Results show linear regression with natural log(fold-change in mRNA expression) as the dependent variables and natural log(viral *NI*) detection as the independent variable. Adjustments for sex and age were not significant for any IFN.

**Table 2c.** IFN Protein Production with SARS-CoV-2 Infection.

| Protein* | Estimate | SE | t | OR | CI | p |
| --- | --- | --- | --- | --- | --- | --- |
| IFN-α2 | -1.08 | 0.428 | -2.53 | 0.338 | 0.146-0.783 | 0.016 |
| IFN-β | -0.0957 | 0.0838 | -1.14 | 0.909 | 0.771-1.07 | 0.26 |
| IFN-γ | -0.672 | 0.222 | -3.02 | 0.511 | 0.33-0.79 | 0.004 |
| IFN-λ1 | -0.0813 | 0.11 | -0.74 | 0.922 | 0.743-1.14 | 0.46 |
| IFN-λ2,3† | -0.520 | 0.115 | -4.52 | 0.595 | 0.475-0.745 | <0.001 |

\*Results show log(pg/ml) of each protein as a function of viral detection.
†Adjustment for sex was not significant, but patients had slightly decreased IFN-λ2,3 in response to SARS-CoV-2 infection for each additional year of age (OR = 0.99, 95% CI = 0.988-0.999, $p = 0.048$ ).

**Table 2d.** IFN stimulated gene transcript responses with SARS-CoV-2 Infection.

| mRNA*,† | Estimate | SE | t | OR | CI | p |
| --- | --- | --- | --- | --- | --- | --- |
| *BST-2* * | 3.22 | 1.03 | 3.13 | 25.1 | 3.33-188 | 0.011 |
| *IFIT-1* | 2.97 | 0.777 | 3.82 | 19.5 | 4.25-89.2 | 0.003 |
| *IFIT-3* | 5.5 | 1.43 | 3.86 | 245 | 15-4020 | 0.003 |
| *IFITM-1* | 0.601 | 0.921 | 0.653 | 1.82 | 0.3-11.1 | 0.53 |
| *MX-1* | 1.2 | 0.427 | 2.82 | 3.33 | 1.44-7.7 | 0.018 |

\*Results show log(fold change of each mRNA) as a function of viral detection.
†Adjustments for age and sex were not significant.
\**BST-2* is also known as *Tetherin*.

**Table 2e.** IFN Stimulated Proteins Using Data Independent Acquisition by Mass Spectrometry.

| **Protein\*** | **$\chi^2$** | **p** |
| --- | --- | --- |
| BST-2 | 5.48 | 0.019 |
| IFIT-1 | 8.33 | 0.004 |
| IFIT-3 | 8.33 | 0.004 |
| IFITM-1<sup>†</sup> | — | — |
| MX-1 | 3.375 | 0.066 |

\* $\chi^2$ tests were applied to $2 \times 2$ tables of detection of protein vs detection of SARS-CoV-2 in all cases except for IFIT-1. There was detection of IFIT-1 in nearly all samples, however, they segregated into high level or low level detection, and the $\chi^2$ test was applied to a $2 \times 2$ table of high detection of protein vs detection of SARS-CoV-2. For each result shown, there were n = 6 SARS-CoV-2 negative and n = 6 SARS-CoV-2 positive patients.
<sup>†</sup> Not Detected.

## Discussion

Our results show large increases in transcription of multiple genes involved in innate immune and inflammatory responses soon after SARS-CoV-2 infection and the development of viral-like symptoms (Tables 1 and 2 and Figures 2 and 3). However, there was a broad-based discrepancy in translation response relative to increased transcription signals similar to the host shut off patterns seen with multiple viruses, including human CoVs such as SARS-CoV that have been reported by many and reviewed by others,^70–74^ and that is just beginning to be described in SARS-CoV-2.^75,76^ An alternative possibility is that proteins are rapidly degraded after translation, however, either possibility is detrimental to a fully functional innate immune response.

Among the IFNs that we evaluated, several had large increases in transcription that were also strongly associated with viral load (Table 2a and 2b), but protein production was either unchanged or decreased when comparing samples from symptomatic infected patients to uninfected controls (Table 2c). For pro-inflammatory cytokines, there were similar large increases in transcription (Table 1a) but no change in measured protein production except for IP-10 alone (Table 1b). Considering that our samples were collected soon after initial symptoms from ambulatory patients, the protein production result may indicate that IP-10 is among the first inflammatory proteins to increase early in infection.

The discrepancies between transcription and translation did not fully extend to the ISGs. We selected to evaluate these molecules because of their importance for anti-viral defense.^60–68^ Observed enormous increases in transcription (Table 2d) were accompanied by several large but uncorrelated (Supplemental Table 2) increases in protein production (Table 2e). Three antiviral ISGs had increased transcription and translation: *IFIT-3* most strongly (Figure 4), *IFIT-1* and *Tetherin*, and there was an additional borderline finding for *MX-1* (Table 2e).

In SARS, suppression of anti-viral proteins occurred late in clinical disease,^77^ however, our results suggest that with SARS-CoV-2, it occurs near the beginning of symptoms. Host translation suppression in SARS is associated with spike protein and non-structural protein 1 (NSP1) interactions with eukaryotic initiation factor-(eIF)-3 which is required for protein translation.^72^ Two recent publications investigating mechanisms involving Nsp1 for SARS-CoV-2 showed similar interference with eIF-3.^75,76^ Our results add to the *in vitro* work by demonstrating supportive evidence from early in the clinical course of human infection. Because viruses depend on host mechanisms for translation of viral proteins that are required for assembly of new infectious particles, our observation of continuing transcription and suppressed translation of human proteins may help explain persistent RT-PCR detection of viral RNA but marked decreases in infectious viral particle production soon after the appearance of symptoms. These tentative hypotheses await further development and testing.

*ACE-2* mRNA was increased among patients with SARS-CoV-2 infection. The finding indicates at least two possible causal relationships. SARS-CoV-2 may selectively infect people with existing high levels of *ACE-2* transcription, or infection itself may increase transcription of *ACE-2* above normal. In either case, increased transcription leading to increased protein expression of ACE-2 likely would increase viral entry and thus help amplify viral replication.

We measured transcripts for three intracellular proteins important in pathways leading to IFN production and initiation of *NFκB*-related inflammation. Transcription of *TBK-1* and *STING-1* were unchanged while there was a decrease in *TRAF-1* mRNA. TRAF-1 is involved in several distinct inflammation-related pathways, but a reduction is most likely associated with increased NFκB activity and subsequently increased systemic inflammation.^58^ The other two proteins, TBK-1 and STING-1, are important for transmitting detection of viral invasion to processes that produce anti-viral IFNs.^41^ There was no increase in transcription of *STING-1* and *TBK-1*, however, *IFN-λ1* and *IFN-λ2* transcripts were markedly elevated (Table 2a and Figure 3). The increases in these transcripts were closely associated with viral load. These findings suggest that detection of viral invasion is successful in generating a signal to increase both systemic inflammation and IFN production. Massive and strongly significant increases in *IFN-λ1* and *IFN-λ2* mRNA (Table 2a) may indicate the critical importance of Type III IFNs in SARS-CoV-2^78–80^ even if protein production was decoupled from high levels of gene transcription by the time our samples were collected.

IP-10 was the sole inflammatory cytokine detected with higher protein concentrations in our samples from infected patients (Table 1b). IP-10 promotes inflammation in Human Immunodeficiency Virus,^81^ H5N1 Influenza A,^82,28^ Middle-East Respiratory Syndrome virus^83^ and SARS-CoV^33^ infections, thus its prominence early in SARS-CoV-2 infection, while unsurprising, may be important for understanding evolution of disease from initial mildly symptomatic to severe and sometimes fatal. Nevertheless, we found a moderately strong inverse relationship with age such that 10 or 25 additional years of age seemed to be associated with dampening of increases in IP-10 (see Results). This inverse association is at odds with the clinical observation of worsening disease severity associated with older ages and generates questions about the nature of previously observed detrimental effects of IP-10^14–17^ on morbidity and mortality with SARS-CoV-2 infection. Older individuals with COVID-19, for example, may be more sensitive to suppression of anti-viral defenses while younger individuals are more sensitive to excessive inflammation. The observations and questions show that abnormal transcript and protein responses to infection cannot be fully interpreted without clinical context drawn from evaluation of a larger study population.

In contrast to limited but interesting results with adjustments for age, adjustments for sex were uninformative. The lack of significant findings may be due to survivor biases. Ill and severely ill patients are less likely to be female,^15^ but the susceptibility to infection associated with sex is unknown. Among patients who develop symptoms, innate immune responses may be similar regardless of sex.

Our study is limited by its cross-sectional design, small size and the nature of the nasopharyngeal swab samples. Due to the urgency of need, we obtained deidentified samples quickly in exchange for giving up detailed clinical annotation. We do not yet have sufficient information to interpret observed abnormalities in IFN and systemic inflammation to seek out associations with clinical outcomes such as respiratory failure or death. However, because a random sample of the population visiting our drive-through diagnosis centers will contain predominantly survivors of infection who never require hospitalization, the measurements we report should roughly represent patients who generally suffer non-severe disease.

The small size of the study limits our ability to generalize our interpretations and conclusions. Sensitivity analyses, however, increase our confidence in the stability of our findings and suggest that there are additional multivariable associations between biomarkers, infection status and viral load that may be explored, further strengthening the impression that additional study of more individuals is needed to better understand the extent of innate immune disruptions due to SARS-CoV-2 infection.

Nasopharyngeal swab samples necessarily retrieve a variety of cell types and may retrieve secreted substances that originate elsewhere than the upper airway. Because samples were frozen prior to evaluation, characterization of cell types by cell counting or flow cytometry was not possible. Prospective human study with immediate processing to allow better assessments of cell types present is possible, but full characterization of secreted molecules will require carefully designed cell culture models to prevent inclusion of molecules produced elsewhere and transported to the nasopharynx. Despite the limitations, our study provides information highlighting several areas of IFN and inflammatory biology that deserve future investigation.

Although our study identifies strong IFN and systemic inflammatory signal transcription responses to infection, only a larger prospective study incorporating careful annotation of patient characteristics, analysis of serial samples with disease progression and reporting of outcomes can fully assess the clinical implications of these initial findings. Our results overall, even with a small study size, emphasize that there are remarkable disruptions early in disease in the immune landscape. Further study is likely to be both fruitful and illuminating.

## Methods

### Samples and Study Population

Our project was reviewed at the University of Utah by both the Institutional Review Board and the Biosafety Committee. An exemption from informed consent was allowed because patient samples were de-identified. All samples were handled in a biosafety level (BSL) 2 capable hood (ThermoFisher Scientific, Waltham, MA, USA) using BSL 3 procedures until virus inactivation and were handled with BSL 2 procedures thereafter.

Randomly selected and completely deidentified, residual nasopharyngeal swab samples from patients presenting for diagnosis of symptoms consistent with COVID-19 during the period of late April through early June of 2020 were enrolled in the study. Clinical testing involved use of a portion of each sample to test with automated, FDA Emergency Use Authorized RT-PCR or transcription-mediated amplification tests for qualitative presence of SARS-CoV-2 RNA. We received sample remainders annotated with age, sex and qualitative nucleic acid amplification-detection results after being frozen at −80 °C for approximately one month.

### Initial Extraction of Human RNA and Proteins

We extracted RNA using Chemagic reagents and Chemagic MSM I extraction platform (Perkin-Elmer, Billerica, MA, USA) from part of each sample remainder producing sufficient RNA to allow real-time polymerase chain reaction (RT-PCR) measurement of reference gene *Pol2A* (mean *C*_*T*_ = 32.22, SD = 4.86). For all other mRNA measurements, we used the Pol2A *C*_*T*_ as the reference point for each individual to calculate fold change (see Methods for *C*_*T*_ definition and usage).

Protease inhibitor cocktail and equal volume of Hank’s Balanced Salt solution (Sigma Aldrich) were added to the final portion of the thawed patient samples prior to centrifugation (20,000 g for 20 min at 4 °C). We carefully aspirated the supernatant for Bead Based Multiplex Immunoassays. Pellets from centrifugation were extracted using All-Prep Micro kits (Qiagen) in accordance with the manufacturer’s instructions, producing additional RNA suitable for RT-PCR and a final protein-containing pellet for Mass Spectrometry.

### Real-Time Polymerase Chain Reaction of viral and human mRNA

For most mRNAs, we had sufficient sample to study all 40 patients; for selected mRNAs, we were able to study 6 samples with and 6 samples without SARS-CoV-2 detection. All specific mRNA measurements were based on RT-PCR employing RNA from a single extraction method to avoid technical sources of noise.

An equal volume of RNA was taken for first strand cDNA reverse transcription (ABI High Capacity cDNA Reverse Transcription Kit) and specific amplification in a StepOnePlus (ABI, ThermoFisher Scientific). Gene specific primers were designed using the Roche Applied Science Universal Probe Library Assay Design Center. All amplifications were performed using a 2-step amplification protocol with ABI PowerUp SYBR Green Master Mix as follows: 1 cycle at 50° C for 2 minutes to activate UDG, 1 cycle at 95° C for 2 minutes to release the DNA polymerase then 40-50 cycles with a 3 second denaturing at 95° C followed by 30 second annealing and denaturing at 60° C.

A melt curve (dissociation) was performed for every primer to ensure the above amplification conditions resulted in the amplification of a single peak. All of the designed primers gave a single peak upon dissociation after amplification suggesting no non-specific binding to other genes. Amplification of genomic DNA was prevented by using primers that spanned an intron. The *IFN-α2* gene and *IFITM-1* and *IFITM-3* ISGs do not have introns. The primers did, however, give a single peak upon dissociation. All other primers including *IFN-λ* spanned an intron.

### Bead Based Multiplex Immunoassays

Cytokine analyses of patient samples were performed using a commercially available enzyme-linked immunosorbent assay (LEGENDplex Human Anti-Virus Response Panel 13-Plex with Filter Plate, BioLegend, San Diego, CA). This bead-based multiplex assay allowed for the simultaneous quantification of interleukins (IL-1β, IL-6, IL-8 [or CXCL8], IL-10, IL-12p70); interferons (IFN-α2, IFN-β, IFN-γ, IFN-λ1, IFN-λ2,3), TNF-α, IP-10, and GM-CSF in patient samples using a flow cytometric approach. All standards and samples were assayed in duplicate using manufacturer recommended protocols. Incubation steps were conducted at room temperature with constant agitation (500 rpm), and shielded from exposure to light. Performing the assay in standard 96-well filter plates facilitated thorough washing of samples and required the use of a MultiScreen Vacuum Manifold (EMD Millipore Corporation, Billerica, MA) alongside a uniform vacuum source. Following the final wash of combined sample and biotinylated detection antibodies, bound proteins of interest were re-suspended in 0.008% final concentration of EM-grade glutaraldehyde (Electron Microscopy Sciences, Hatfield, PA, USA, Cat #16216) for 48 hours at 4° C to inactivate SARS-CoV-2, adapting a protocol previously investigated for SARS-CoV.^84^ Following incubation, samples were washed a final time and transferred to a polystyrene 96-well plate with a conical bottom. Flow cytometric analysis of cytokines was performed using a BD FACSCanto II system (BD Biosciences, San Jose, CA) at the University of Utah Flow Cytometry Core (Salt Lake City, UT) and analyzed using LEGENDplex software (BioLegend).

### Mass Spectrometry

#### Preparation of proteins prior to mass spectrometry

Proteins were reduced with 5 mM dithiothreitol (DTT) at 60° C for 45 minutes, followed by alkylation with 10 mM iodoacetamide (IAA) at room temperature for 30 minutes in the dark. Excess IAA was neutralized by addition of 5 mM DTT. A trypsin/LysC mixture (Promega; Madison, WI) was added to the proteins in a 1:100 ratio and the proteins were digested overnight at 38° C. The digestion was quenched by acidification of the solution with the addition of 1% formic acid to a pH of 2-3.

Initially, the pelleted proteins from the COVID-19 patients would not completely dissolve in the 50 mM ammonium bicarbonate. However, after the trypsin/LysC digestion all of the samples were completely dissolved in solution. The final concentration of the peptides was determined using a peptide colorimetric assay and the use of a Nanodrop One (ThermoFisher Scientific) spectrophotometer.

#### DDA nanoLC-MS/MS

Peptides (1 μg on column) were loaded using a Dionex UltiMate 3000 RSLCnano system (ThermoFisher Scientific) onto a PharmaFluidics μPAC micro-chip based trapping column and separated using a 50 cm equivalent PharmaFluidics μPAC micro-chip based column (PharmaFluidics, Ghent, Belgium). Chromatography was performed using ultrapure water with 0.1% formic acid (solvent A) and acetonitrile containing 0.1% formic acid (solvent B). Elution was carried out with an initial mobile phase concentration of 5% for 4 minutes followed by a ramp to 45% over 76 minutes then a second ramp to 95% B in 5 minutes. This was held for 10 minutes followed by ramping down to 5% B over two minutes and re-equilibration for 10 minutes. Flow rate was 0.5 mL/min. A QExactive HF (ThermoFisher Scientific) coupled to a Flex nano spray source was employed with the following settings for MS1; resolution 60, AGC target 3e6, maximum IT 100 ms, scan range 375-1650 m/z. MS2 settings were; resolution 15,000, AGC target 2e5, maximum IT 25 ms, isolation window 1.4 m/z. Top 15 DDA analysis was performed with NCE set to 27.

#### DIA nanoLC-MS/MS

Staggered window DIA analysis was carried out using the methods described by Pino *et. al*.^85^ A peptide centric gas phase retention time library was generated by pooling equal amounts of each sample and analyzing this using six narrow window DIA experiments with the following settings for MS1: resolution 60,000, AGC target 1e6, maximum IT 55, with 6 separate analyses in the following mass ranges 395-505 m/z, 495-605 m/z, 595-705 m/z, 695-805 m/z, 795-905 m/z, and 895-1005 m/z. MS2 analysis used the following settings: resolution 30,000, AGC target 1e6, loop count 25, default charge 3, NCE 27 with 4 m/z staggered DIA windows. NanoLC-MS/MS analysis was carried out identically to DDA analysis described above.

#### DDA data processing

The Proteome Discover version 2.4 (ThermoFisher Scientific) precursor-based quantification processing workflow was employed. SequestHT with multiple peptide search and percolator validation was employed to extract protein data. The following search options were employed, *Homo sapiens* fasta file, trypsin digestion, 2 missed cleavages, minimum peptide length 6, precursor mass tolerance 10 ppm, fragment mass tolerance 0.02 Da, carbamidomethylation of cysteine as a peptide static modification, N-terminal acetylation, N-terminal Met-loss and methionine oxidation as protein dynamic modifications.

#### DIA data processing

Thermo .RAW files were demultiplexed and converted to mzML files using MSConvert.^86^ The Walnut functionality of EnclopeDIA^87^ was employed for peptide centric library creation. Peptides were identified using the same variables as DDA described above. Quantitation was performed using Skyline.^88^

#### Calculations and Statistical Analysis

For all mRNA, we calculated fold-change for each sample (*FCsample*) after measuring the fractional number of polymerase chain reaction doubling cycles required so that SYBR Green fluorescence exceeded the threshold for detection (*C*_*T*_). We used the following formula:

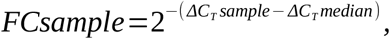

where *ΔC*_*T*_*sample* was the number of doubling cycles to detect each mRNA minus the number of doubling cycles to detect mRNA from the *Pol2A* reference gene for each sample, and the *ΔC*_*T*_*median* was the median *ΔC*_*T*_*sample* for samples without detection of SARS-CoV-2. The incorporation of *C*_*T*_ for *Pol2A* mRNA in the calculation indexes the measurement so that samples with different efficiencies of recovery of mRNA containing cells between testing individuals are standardized.

For IFIT-3 fold change values from DDA mass spectrometry, *FCsample* was calculated:

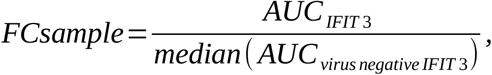

where *AUC*_*IFIT3*_ is the area under the curve (AUC) for peptides identified as part of IFIT-3, and *median*(*AUC*_*virus negative IFIT3*_) is the median AUC for IFIT-3 from samples without detection of SARS-CoV-2. The other pre-specified ISG proteins were undetectable by DDA mass spectrometry, and thus no fold-change calculation was possible.

We calculated summary statistics. We examined associations between different biomarker measurements by calculating Spearman’s rank correlation statistic to better understand potential dependencies. We used log transformations of all mRNA and protein measurements in our statistical calculations because of the log-normal nature of our results. Others using the methods that we employed, however, often report results using either natural or base 2 logs. Because the bulk of our results are commonly reported using natural log values, we standardized on those for reporting. The effect is to slightly change results normally reported using base 2 logs by a proportion equal to natural log of 2 (or 0.698). This usage has no effect on interpretations of results.

Using SARS-CoV-2 infection status as the independent variable, we performed linear regression with natural log(*FC*) of each pre-specified mRNA or natural log(*FC*_*IFIT3*_) as the dependent variable because we seek to understand the biological effects of infection. Each univariable model was adjusted with age and sex with backward selection to understand the impacts on model fits. We performed univariable linear regression with natural log(*FC*) or natural log(protein concentration [pg/ml]) as the dependent variables and natural log(*FC*_*viral N1 protein mRNA*_) as the independent variable to understand associations with viral load, adjusting with age and sex as above.

We performed sensitivity analyses of significant associations reported in Tables 1 and 2. For each dependent biomarker with significant associations with infection status or viral load, we selected all other biomarkers reported to have significant correlations in Supplementary Table 2 as additional adjustment variables. Using these adjustment biomarkers one at a time, we assessed the impact on the estimates for infection status and viral load for each significant association reported in Tables 1 and 2.

We assigned 50 as the *C*_*T*_ value for undetectable mRNA. For undetectable proteins by bead based multiplex immunoassay, we assigned the minimum detection value. These assignments enable quantitative analysis without treating the values as missing. Results were similar when analysis was restricted to raw data derived from the 6 infected and 6 uninfected samples with the highest recovery of RNA. All calculations and statistical modeling were performed using the R statistical system.^89^

## Data Availability

The final data set used to generate all results is available upon request to the corresponding author. Samples are not available as most were consumed by the studies presented.

## Acknowledgments

We recognize and thank the many people at the University of Utah who have faced blistering heat, freezing cold, earthquakes, hurricane force winds, forest fire smoke and the threat of infection to consistently obtain high quality diagnostic samples in service to the community which also supplied the material for our study.

This study was funded by the Ben B and Iris M Margolis Family Foundation of Utah and the Claudia Ruth Goodrich Stevens Endowment Fund at the University of Utah. This work was supported by the University of Utah Flow Cytometry Facility in addition to the National Cancer Institute through Award Number 5P30CA042014-24. Proteomics mass spectrometry analysis was performed at the Mass Spectrometry and Proteomics Core Facility at the University of Utah which is supported by U54 DK110858 05. Mass spectrometry equipment was obtained through a Shared Instrumentation Grant 1 S10 OD018210 01A1. Oligonucleotide primers were synthesized by the DNA/Peptide Facility, part of the Health Sciences Center Cores at the University of Utah.

## Author Contributions

TGL and FRA initiated the project. TGL, FRA, DRC, JEC, MNH, NDH, JLJ, CK, DTL, KAP, ABS, KJW, LJWa, LJWe and RPIII designed the study. SCH oversaw the drive-through tent clinical testing program. TGL, KEH, SCH, DTL, JLJ, SMS and KT obtained samples. ABS and EZ performed RT-PCR. GJG, MNH, YL and JEM performed bead-based multianalyte immunoassays. MNH, SMS, YL and KT extracted RNA. JEC, KT, MNH, SMOS and YL prepared proteins for mass spectrometry. JEC and SMOS performed mass spectrometry. TGL, FRA, DRC, JEC and CK analyzed the data. TGL initiated the manuscript writing, but all authors, FRA, BCC, DRC, JEC, GJG, KEH, SCH, NDH, MNH, JLJ, CK, YL, DTL, JEM, EAM, SMOS, KAP, SMS, ABS, KT, KJW, LJWa, LJWe, EZ and RPIII participated in writing, formulation of interpretations and in revisions. TGL, JEC, JEM and RPIII obtained funding. JLJ, KAP, LJWa and LJWe managed regulatory aspects of the study. Authors are listed in alphabetical order except first and senior (last) authors.

## Competing Interests

TGL, JLS and KAP are supported by NIH/NHLBI R01 125520 and by the US Cystic Fibrosis Foundation, Bethesda, MD, grants LIOU13A0, LIOU14P0, LIOU14Y4, LIOU15Y4. During the course of the study, TGL, JLS and KAP received other support for performing cystic fibrosis-related clinical trials from Abbvie, Inc; Corbus Pharmaceuticals Holdings, Inc; Gilead Sciences, Inc.; Laurent Pharmaceuticals, Inc; Nivalis Therapeutics, Inc; Novartis Pharmaceuticals; Proteostasis Therapeutics, Inc; Savara, Inc; and Vertex Pharmaceuticals, Inc. None of the funding was provided for development of vaccines or treatments for SARS-CoV-2 infection. FRA received other support from the National Science Foundation (grant EMSW21-RTG). JEC is supported by NIH/NIDDK U54 DK110858 05. MNH is supported by NIH/NHLBI R01HL137033. CK received support from the UK Medical Research Council. DTL is supported by NIH/NAIAD R01AI130378. The sponsors of clinical trials and funders of other support played no roles in the current study.

## Notes

### Author Declarations

Our project was reviewed at the University of Utah by both the Institutional Review Board and the Biosafety Committee. An exemption from informed consent was allowed because patient samples were de-identified.

## References

1. Timeline: WHO’s COVID-19 response. https://www.who.int/emergencies/diseases/novel-coronavirus-2019/interactive-timeline.

2. Lu, R. et al. Genomic characterisation and epidemiology of 2019 novel coronavirus: implications for virus origins and receptor binding. Lancet 395, 565–574 (2020).

3. Wu, F. et al. A new coronavirus associated with human respiratory disease in China. Nature 579, 265–269 (2020).

4. Corman, V. M. et al. Detection of 2019 novel coronavirus (2019-nCoV) by real-time RT-PCR. Euro Surveill. 25, (2020).

5. Cheng, H.-Y. et al. Contact Tracing Assessment of COVID-19 Transmission Dynamics in Taiwan and Risk at Different Exposure Periods Before and After Symptom Onset. JAMA Intern Med (2020) doi:10.1001/jamainternmed.2020.2020.

6. Kang, J. et al. South Korea’s responses to stop the COVID-19 pandemic. Am J Infect Control 48, 1080–1086 (2020).

7. Wang, C. J., Ng, C. Y. & Brook, R. H. Response to COVID-19 in Taiwan: Big Data Analytics, New Technology, and Proactive Testing. JAMA (2020) doi:10.1001/jama.2020.3151.

8. Cowling, B. J. et al. Impact assessment of non-pharmaceutical interventions against coronavirus disease 2019 and influenza in Hong Kong: an observational study. Lancet Public Health 5, e279–e288 (2020).

9. Baker, M. G., Kvalsvig, A. & Verrall, A. J. New Zealand’s COVID-19 elimination strategy. Medical Journal of Australia 213, 198-200.e1 (2020).

10. Fouda, A., Mahmoudi, N., Moy, N. & Paolucci, F. The COVID-19 pandemic in Greece, Iceland, New Zealand, and Singapore: Health Policies and Lessons Learned. Health Policy Technol (2020) doi:10.1016/j.hlpt.2020.08.015.

11. Sakurai, A. et al. Natural History of Asymptomatic SARS-CoV-2 Infection. N Engl J Med 383, 885–886 (2020).

12. Wölfel, R. et al. Virological assessment of hospitalized patients with COVID-2019. Nature 581, 465– 469 (2020).

13. Gandhi, M., Beyrer, C. & Goosby, E. Masks Do More Than Protect Others During COVID-19: Reducing the Inoculum of SARS-CoV-2 to Protect the Wearer. J Gen Intern Med (2020) doi:10.1007/s11606-020-06067-8.

14. Zhou, P. et al. A pneumonia outbreak associated with a new coronavirus of probable bat origin. Nature 579, 270–273 (2020).

15. Huang, C. et al. Clinical features of patients infected with 2019 novel coronavirus in Wuhan, China. Lancet 395, 497–506 (2020).

16. Lescure, F.-X. et al. Clinical and virological data of the first cases of COVID-19 in Europe: a case series. Lancet Infect Dis 20, 697–706 (2020).

17. Wang, D. et al. Clinical Characteristics of 138 Hospitalized Patients With 2019 Novel Coronavirus– Infected Pneumonia in Wuhan, China. JAMA 323, 1061–1069 (2020).

18. Middleton, E. A. et al. Neutrophil Extracellular Traps (NETs) Contribute to Immunothrombosis in COVID-19 Acute Respiratory Distress Syndrome. Blood (2020) doi:10.1182/blood.2020007008.

19. Manne, B. K. et al. Platelet Gene Expression and Function in COVID-19 Patients. Blood (2020) doi:10.1182/blood.2020007214.

20. Chen, G. et al. Clinical and immunological features of severe and moderate coronavirus disease 2019. J. Clin. Invest. 130, 2620–2629 (2020).

21. Zhou, Z. et al. Heightened Innate Immune Responses in the Respiratory Tract of COVID-19 Patients. Cell Host Microbe (2020) doi:10.1016/j.chom.2020.04.017.

22. Ye, Q., Wang, B. & Mao, J. The pathogenesis and treatment of the ‘Cytokine Storm’ in COVID-19. J. Infect. (2020) doi:10.1016/j.jinf.2020.03.037.

23. Wang, Z. et al. Early hypercytokinemia is associated with interferon-induced transmembrane protein-3 dysfunction and predictive of fatal H7N9 infection. Proc. Natl. Acad. Sci. U.S.A. 111, 769–774 (2014).

24. Pedersen, S. F. & Ho, Y.-C. SARS-CoV-2: A Storm is Raging. J. Clin. Invest. (2020) doi:10.1172/JCI137647.

25. Long, Q.-X. et al. Clinical and immunological assessment of asymptomatic SARS-CoV-2 infections. Nat. Med. 26, 1200–1204 (2020).

26. Zhang, X. et al. Viral and host factors related to the clinical outcome of COVID-19. Nature 1–4 (2020) doi:10.1038/s41586-020-2355-0.

27. Galani, I. E. et al. Interferon-λ Mediates Non-redundant Front-Line Antiviral Protection against Influenza Virus Infection without Compromising Host Fitness. Immunity 46, 875-890.e6 (2017).

28. de Jong, M. D. et al. Fatal outcome of human influenza A (H5N1) is associated with high viral load and hypercytokinemia. Nat Med 12, 1203–1207 (2006).

29. Gralinski, L. E. & Baric, R. S. Molecular pathology of emerging coronavirus infections. J. Pathol. 235, 185–195 (2015).

30. Kim, E. S. et al. Clinical Progression and Cytokine Profiles of Middle East Respiratory Syndrome Coronavirus Infection. J. Korean Med. Sci. 31, 1717–1725 (2016).

31. Faure, E. et al. Distinct immune response in two MERS-CoV-infected patients: can we go from bench to bedside? PLoS ONE 9, e88716 (2014).

32. Zhou, B. et al. The nucleocapsid protein of severe acute respiratory syndrome coronavirus inhibits cell cytokinesis and proliferation by interacting with translation elongation factor 1alpha. J. Virol. 82, 6962–6971 (2008).

33. Chen, J. & Subbarao, K. The Immunobiology of SARS*. Annu. Rev. Immunol. 25, 443–472 (2007).

34. Spiegel, M. et al. Inhibition of Beta interferon induction by severe acute respiratory syndrome coronavirus suggests a two-step model for activation of interferon regulatory factor 3. J. Virol. 79, 2079– 2086 (2005).

35. Qian, Z. et al. Innate immune response of human alveolar type II cells infected with severe acute respiratory syndrome-coronavirus. Am. J. Respir. Cell Mol. Biol. 48, 742–748 (2013).

36. Totura, A. L. & Baric, R. S. SARS coronavirus pathogenesis: host innate immune responses and viral antagonism of interferon. Current Opinion in Virology 2, 264–275 (2012).

37. Nicholls, J. M. et al. Lung pathology of fatal severe acute respiratory syndrome. Lancet 361, 1773–1778 (2003).

38. Zielecki, F. et al. Human cell tropism and innate immune system interactions of human respiratory coronavirus EMC compared to those of severe acute respiratory syndrome coronavirus. J. Virol. 87, 5300– 5304 (2013).

39. Kindler, E. et al. Efficient replication of the novel human betacoronavirus EMC on primary human epithelium highlights its zoonotic potential. mBio 4, e00611–00612 (2013).

40. Li, S.-W. & Lin, C.-W. Human coronaviruses: Clinical features and phylogenetic analysis. Biomedicine (Taipei) 3, 43–50 (2013).

41. Lei, J. & Hilgenfeld, R. RNA-virus proteases counteracting host innate immunity. FEBS Lett. 591, 3190–3210 (2017).

42. Nelemans, T. & Kikkert, M. Viral Innate Immune Evasion and the Pathogenesis of Emerging RNA Virus Infections. Viruses 11, 961 (2019).

43. Lazear, H. M., Schoggins, J. W. & Diamond, M. S. Shared and Distinct Functions of Type I and Type III Interferons. Immunity 50, 907–923 (2019).

44. DeDiego, M. L. et al. Coronavirus virulence genes with main focus on SARS-CoV envelope gene. Virus Res. 194, 124–137 (2014).

45. Ziegler, T. et al. Severe Acute Respiratory Syndrome Coronavirus Fails To Activate Cytokine-Mediated Innate Immune Responses in Cultured Human Monocyte-Derived Dendritic Cells. Journal of Virology 79, 13800–13805 (2005).

46. Spiegel, M. & Weber, F. Inhibition of cytokine gene expression and induction of chemokine genes in non-lymphatic cells infected with SARS coronavirus. Virology Journal 3, 17 (2006).

47. Deonarain, R. et al. Impaired antiviral response and alpha/beta interferon induction in mice lacking beta interferon. J Virol 74, 3404–3409 (2000).

48. Hoffmann, M. et al. SARS-CoV-2 Cell Entry Depends on ACE2 and TMPRSS2 and Is Blocked by a Clinically Proven Protease Inhibitor. Cell 181, 271-280.e8 (2020).

49. Heurich, A. et al. TMPRSS2 and ADAM17 cleave ACE2 differentially and only proteolysis by TMPRSS2 augments entry driven by the severe acute respiratory syndrome coronavirus spike protein. J. Virol. 88, 1293–1307 (2014).

50. Matsuyama, S., Ujike, M., Morikawa, S., Tashiro, M. & Taguchi, F. Protease-mediated enhancement of severe acute respiratory syndrome coronavirus infection. Proc. Natl. Acad. Sci. U.S.A. 102, 12543–12547 (2005).

51. Haagmans, B. L. et al. Pegylated interferon-alpha protects type 1 pneumocytes against SARS coronavirus infection in macaques. Nat. Med. 10, 290–293 (2004).

52. Cinatl, J. et al. Treatment of SARS with human interferons. Lancet 362, 293–294 (2003).

53. Mantlo, E., Bukreyeva, N., Maruyama, J., Paessler, S. & Huang, C. Antiviral activities of type I interferons to SARS-CoV-2 infection. Antiviral Res. 179, 104811 (2020).

54. Cox, D. R. Planning of Experiments. (Wiley, 1958).

55. Udugama, B. et al. Diagnosing COVID-19: The Disease and Tools for Detection. ACS Nano 14, 3822– 3835 (2020).

56. Lu, X. et al. US CDC Real-Time Reverse Transcription PCR Panel for Detection of Severe Acute Respiratory Syndrome Coronavirus 2. Emerg Infect Dis 26, (2020).

57. Shang, J. et al. Cell entry mechanisms of SARS-CoV-2. Proc. Natl. Acad. Sci. U.S.A. (2020) doi:10.1073/pnas.2003138117.

58. Lalani, A. I., Zhu, S., Gokhale, S., Jin, J. & Xie, P. TRAF molecules in inflammation and inflammatory diseases. Current Pharmacology Reports 4, 64–90 (2018).

59. Edilova, M. I., Abdul-Sater, A. A. & Watts, T. H. TRAF1 Signaling in Human Health and Disease. Front Immunol 9, 2969 (2018).

60. Sadler, A. J. & Williams, B. R. G. Interferon-inducible antiviral effectors. Nat. Rev. Immunol. 8, 559– 568 (2008).

61. Haller, O., Staeheli, P., Schwemmle, M. & Kochs, G. Mx GTPases: dynamin-like antiviral machines of innate immunity. Trends Microbiol. 23, 154–163 (2015).

62. Verhelst, J., Hulpiau, P. & Saelens, X. Mx proteins: Antiviral gatekeepers that restrain the uninvited. Microbiology and Molecular Biology Reviews 77, 551–566 (2013).

63. Kochs, G., Janzen, C., Hohenberg, H. & Haller, O. Antivirally active MxA protein sequesters La Crosse virus nucleocapsid protein into perinuclear complexes. Proc. Natl. Acad. Sci. U.S.A. 99, 3153–3158 (2002).

64. Hefti, H. P. et al. Human MxA protein protects mice lacking a functional alpha/beta interferon system against La crosse virus and other lethal viral infections. J. Virol. 73, 6984–6991 (1999).

65. Kochs, G. & Haller, O. Interferon-induced human MxA GTPase blocks nuclear import of Thogoto virus nucleocapsids. Proc. Natl. Acad. Sci. U.S.A. 96, 2082–2086 (1999).

66. Diamond, M. S. & Farzan, M. The broad-spectrum antiviral functions of IFIT and IFITM proteins. Nat Rev Immunol 13, 46–57 (2013).

67. Perreira, J. M., Chin, C. R., Feeley, E. M. & Brass, A. L. IFITMs restrict the replication of multiple pathogenic viruses. J. Mol. Biol. 425, 4937–4955 (2013).

68. Blanco-Melo, D., Venkatesh, S. & Bieniasz, P. D. Origins and Evolution of tetherin, an Orphan Antiviral Gene. Cell Host Microbe 20, 189–201 (2016).

69. Wang, S.-M., Huang, K.-J. & Wang, C.-T. Severe acute respiratory syndrome coronavirus spike protein counteracts BST2-mediated restriction of virus-like particle release. J. Med. Virol. 91, 1743–1750 (2019).

70. Walsh, D., Mathews, M. B. & Mohr, I. Tinkering with translation: protein synthesis in virus-infected cells. Cold Spring Harb Perspect Biol 5, a012351 (2013).

71. Kikkert, M. Innate Immune Evasion by Human Respiratory RNA Viruses. J Innate Immun 12, 4–20 (2020).

72. Xiao, H., Xu, L. H., Yamada, Y. & Liu, D. X. Coronavirus spike protein inhibits host cell translation by interaction with eIF3f. PLoS One 3, e1494 (2008).

73. Kamitani, W., Huang, C., Narayanan, K., Lokugamage, K. G. & Makino, S. A two-pronged strategy to suppress host protein synthesis by SARS coronavirus Nsp1 protein. Nat Struct Mol Biol 16, 1134–1140 (2009).

74. Narayanan, K. et al. Severe acute respiratory syndrome coronavirus nsp1 suppresses host gene expression, including that of type I interferon, in infected cells. J Virol 82, 4471–4479 (2008).

75. Schubert, K. et al. SARS-CoV-2 Nsp1 binds the ribosomal mRNA channel to inhibit translation. Nature Structural & Molecular Biology 27, 959–966 (2020).

76. Thoms, M. et al. Structural basis for translational shutdown and immune evasion by the Nsp1 protein of SARS-CoV-2. Science 369, 1249–1255 (2020).

77. Cheung, C. Y. et al. Cytokine responses in severe acute respiratory syndrome coronavirus-infected macrophages in vitro: Possible relevance to pathogenesis. Journal of Virology 79, 7819–7826 (2005).

78. Andreakos, E. & Tsiodras, S. COVID-19: lambda interferon against viral load and hyperinflammation. EMBO Mol Med (2020) doi:10.15252/emmm.202012465.

79. Jewell, N. A. et al. Lambda interferon is the predominant interferon induced by influenza A virus infection in vivo. J. Virol. 84, 11515–11522 (2010).

80. Davidson, S. et al. IFNλ is a potent anti-influenza therapeutic without the inflammatory side effects of IFNα treatment. EMBO Mol Med 8, 1099–1112 (2016).

81. Lei, J., Yin, X., Shang, H. & Jiang, Y. IP-10 is highly involved in HIV infection. Cytokine 115, 97–103 (2019).

82. Chan, M. C. W. et al. Proinflammatory cytokine responses induced by influenza A (H5N1) viruses in primary human alveolar and bronchial epithelial cells. Respir. Res. 6, 135 (2005).

83. Chu, H. et al. Productive replication of Middle East respiratory syndrome coronavirus in monocyte-derived dendritic cells modulates innate immune response. Virology 454–455, 197–205 (2014).

84. Darnell, M. E. R., Subbarao, K., Feinstone, S. M. & Taylor, D. R. Inactivation of the coronavirus that induces severe acute respiratory syndrome, SARS-CoV. J. Virol. Methods 121, 85–91 (2004).

85. Pino, L. K., Just, S. C., MacCoss, M. J. & Searle, B. C. Acquiring and Analyzing Data Independent Acquisition Proteomics Experiments without Spectrum Libraries. Mol Cell Proteomics 19, 1088–1103 (2020).

86. Chambers, M. C. et al. A cross-platform toolkit for mass spectrometry and proteomics. Nat Biotechnol 30, 918–920 (2012).

87. Searle, B. C. et al. Chromatogram libraries improve peptide detection and quantification by data independent acquisition mass spectrometry. Nat Commun 9, 5128 (2018).

88. MacLean, B. et al. Skyline: an open source document editor for creating and analyzing targeted proteomics experiments. Bioinformatics 26, 966–968 (2010).

89. R Core Team. R: A language and environment for statistical computing. (R Foundation for Statistical Computing, 2020).

