## Supplementary Table 1 for "SARS-CoV-2 Innate Effector Associations and Viral Load in Early Nasopharyngeal Infection"

**Supplementary Table 1a. Demographics**

| Sex | Age Range and SARS-CoV-2 Status | | n |
|---|---|---|---|
| | Not Detected | Detected | |
| male | 11-90 | 24-63 | 23 |
| female | 22-78 | 16-80 | 17 |
| n | 20 | 20 | 40 |

**Supplementary Table 1b. Statistics***

| Age Effect | Estimate | SE | *t* | OR | 95% CI | *p* |
|---|---|---|---|---|---|---|
| Female | -0.0361 | 0.0177 | -2.04 | 0.965 | 0.932-0.999 | 0.041 |
| SARS2 Detected | -0.0365 | 0.0174 | -2.09 | 0.964 | 0.932-0.998 | 0.036 |

*Univariable logistic regression models of age as the input variable with female sex and positive virus detection as output variables. Younger patients are more likely to be female and more likely to be infected.

**Supplementary Table 1c. Relationship of Age with Viral Load by Sex***

| Sex | Estimate | SE | *t* | OR | 95% CI | *p* |
|---|---|---|---|---|---|---|
| Male | -1.22 | 0.39 | -3.14 | 0.294 | 0.137-0.631 | 0.005† |
| Female | 0.195 | 0.482 | 0.404 | 1.21 | 0.472-3.12 | 0.692 |

*Univariable linear regressions between Age in years as the dependent variable and log(FC) of viral N1 protein for each sex.

† There was a strong linear relationship among male patients between decreasing age and increasing viral load but no significant relationship among female patients. There are at least several potential explanations. First, this may be an accident of our request for remainders from clinical testing for SARS-CoV-2 and the small size of the study. The result must be reproduced in a larger sample population before making a firm conclusion despite the apparent strength of the statistical testing. Second, if this result is reproducible, there may be a survivor bias such that older male patients are more likely to bypass outpatient testing because they seek hospitalization or die early. Symptoms prompting diagnostic testing may begin at earlier time points in disease among older males, or viral replication may be less efficient among these individuals or conversely have already progressed down the airway away from the nasopharynx sampling site. Ascertainment of any of these or other possibilities requires additional clinical information not available with these samples.
