## Supplementary Table 2 for "SARS-CoV-2 Innate Effector Associations and Viral Load in Early Nasopharyngeal Infection"

**Supplementary Table 2a. Correlations among measured protein biomarkers.***

| Protein, log(pg/ml) | GMCSF | IFNα2 | IFNβ | IFNγ | IFNλ1 | IFNλ23 | IL1β | IL6 | IL8 | IL10 | IL12p70 | IP10 |
|---|---|---|---|---|---|---|---|---|---|---|---|---|
| **IFNα2** | - | | | | | | | | | | | |
| **IFNβ** | 0.33 | **0.7** | | | | | | | | | | |
| **IFNγ** | 0.35 | **0.72** | **0.62** | | | | | | | | | |
| **IFNλ1** | - | - | - | - | | | | | | | | |
| **IFNλ23** | 0.33 | **0.53** | 0.46 | **0.63** | - | | | | | | | |
| **IL1β** | - | **0.61** | 0.35 | 0.42 | - | 0.36 | | | | | | |
| **IL6** | - | - | 0.32 | - | - | - | - | | | | | |
| **IL8** | - | - | - | - | - | - | - | 0.4 | | | | |
| **IL10** | - | - | - | - | - | - | - | 0.37 | - | | | |
| **IL12p70** | -0.33 | - | - | - | **0.52** | - | - | - | - | 0.45 | | |
| **IP10** | - | - | - | - | - | - | - | **0.57** | 0.38 | 0.4 | - | |
| **TNFα** | - | - | - | - | - | - | - | - | - | - | - | - |
| | **Protein, log(pg/ml)** | | | | | | | | | | | |

**Supplementary Table 2b. Correlations between measured Proteins and mRNA biomarkers.***

| MRNA, log(Fold-Change) | GMCSF | IFNα2 | IFNβ | IFNγ | IFNλ1 | IFNλ23 | IL1β | IL6 | IL8 | IL10 | IL12p70 | IP10 | TNFα |
|---|---|---|---|---|---|---|---|---|---|---|---|---|---|
| ***GMCSF*** | - | - | - | - | - | - | - | - | - | - | - | - | - |
| ***IFNα2*** | - | - | - | - | 0.32 | - | - | - | - | - | 0.33 | - | - |
| ***IFNβ1*** | - | - | - | - | - | - | - | - | - | - | 0.33 | - | - |
| ***IFNγ*** | - | - | - | - | - | - | - | - | - | - | - | - | - |
| ***IFNλ1*** | - | - | - | - | - | - | - | - | - | - | - | 0.49 | - |
| ***IFNλ2*** | - | - | - | - | - | - | - | - | - | - | - | 0.36 | - |
| ***IFNλ3*** | - | - | - | - | - | - | - | - | - | - | - | - | - |
| ***IL6*** | - | - | - | - | - | - | - | - | - | - | - | -0.34 | - |
| ***IL8*** | - | - | - | - | - | - | - | - | - | - | - | - | - |
| ***IL10*** | - | -0.5 | -0.44 | -0.46 | - | **-0.65** | - | - | - | - | - | - | - |
| ***IP10*** | - | - | - | -0.35 | - | -0.32 | - | 0.45 | - | - | - | **0.61** | - |
| ***TNFα*** | - | -0.37 | - | -0.33 | - | **-0.54** | - | - | - | - | - | - | - |
| | **Protein, log(pg/ml)** | | | | | | | | | | | | |

**Supplementary Table 2c. Correlations among measured mRNA biomarkers.***

| mRNA, log(Fold-Change) | *GMCSF* | *IFNα2* | *IFNβ1* | *IFNγ* | *IFNλ1* | *IFNλ2* | *IFNλ3* | *IL6* | *IL8* | *IL10* | *IP10* |
|---|---|---|---|---|---|---|---|---|---|---|---|
| ***IFNα2*** | - | | | | | | | | | | |
| ***IFNβ1*** | - | **1.00** | | | | | | | | | |
| ***IFNγ*** | 0.33 | **0.52** | **0.5** | | | | | | | | |
| ***IFNλ1*** | - | **0.52** | **0.52** | 0.45 | | | | | | | |
| ***IFNλ2*** | - | **0.53** | **0.52** | **0.59** | **0.86** | | | | | | |
| ***IFNλ3*** | - | 0.44 | 0.44 | 0.47 | **0.6** | **0.76** | | | | | |
| ***IL6*** | **0.55** | **0.6** | **0.59** | **0.45** | - | 0.34 | **0.53** | | | | |
| ***IL8*** | 0.4 | 0.4 | 0.39 | 0.33 | - | - | - | - | | | |
| ***IL10*** | 0.48 | - | - | 0.4 | - | 0.33 | - | - | 0.33 | | |
| ***IP10*** | - | - | - | - | 0.34 | 0.36 | - | - | **0.6** | 0.5 | |
| ***TNFα*** | 0.43 | - | - | - | - | - | - | - | 0.38 | **0.77** | 0.47 |
| | **MRNA, log(Fold-Change)** | | | | | | | | | | |

* Spearman Correlations with $p < 0.001$ shown in **Bold Type**, $0.001 \leq p < 0.01$ in Regular Type and $0.01 \leq p < 0.05$ in Grey Type, with $p \geq 0.05$ not shown. Dashes mark non-significant results. Grey Boxes mark self-correlations or duplicate conditions.
