## Supplementary Figure Legends for "SARS-CoV-2 Innate Effector Associations and Viral Load in Early Nasopharyngeal Infection"

**Supplementary Figure 1. Relationships with Age.** In **a.** male patients were older and in **b.** patients without infection were older, but in **c.** this relationship is seen to be limited to male patients (see also Supplementary Table 1c).

**Supplementary Figure 2. SARS-CoV-2 Infection and Signaling Protein Transcription.** There was no relationship between *TBK1* mRNA transcription and **a.** infection status or **b.** viral *N1* protein transcripts. There was also no relationship between *STING1* mRNA transcription and **c.** infection status or **d.** viral *N1* protein mRNA.
