## Supplementary figures and images for "SARS-CoV-2 Innate Effector Associations and Viral Load in Early Nasopharyngeal Infection"

### Supplementary Figure 1

#### Supplementary Figure 1

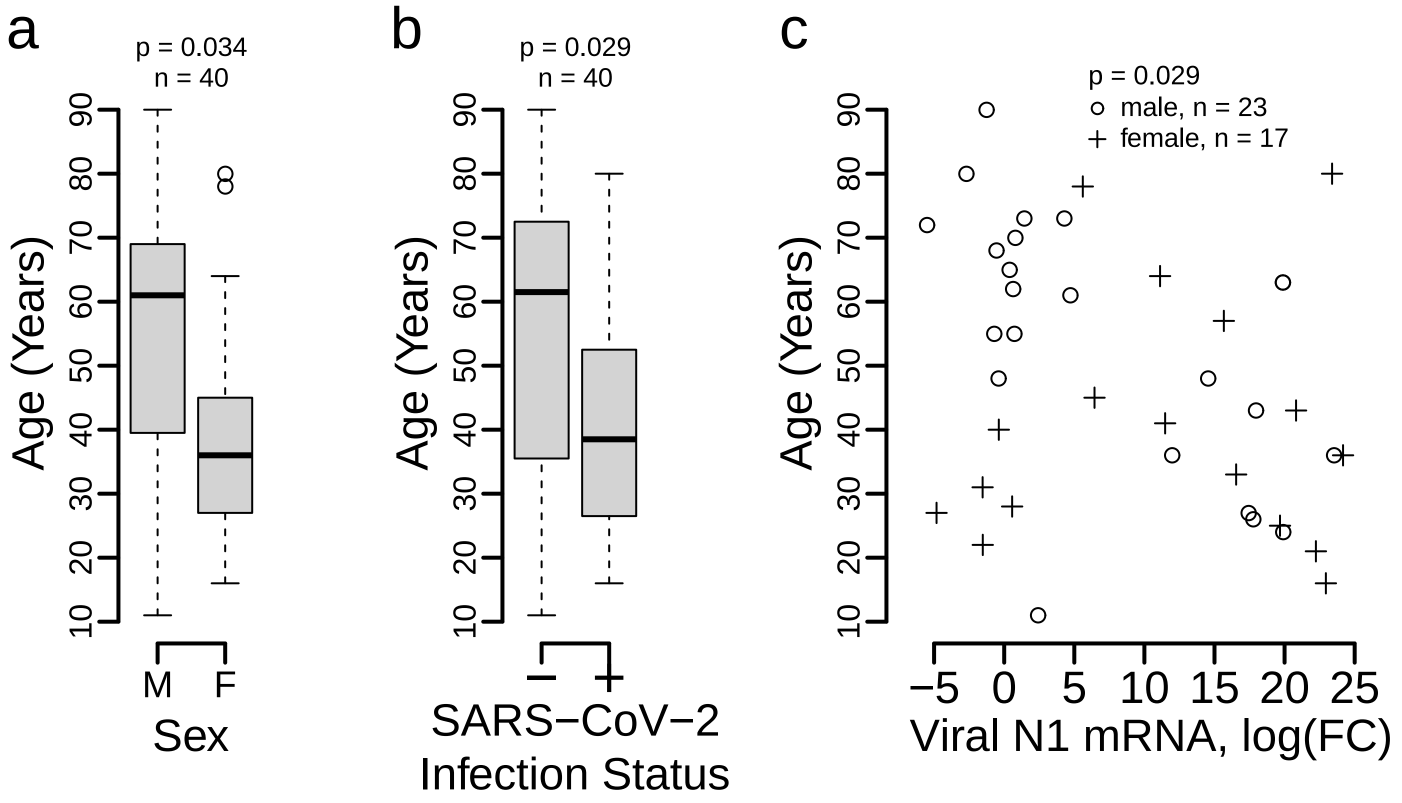

### Supplementary Figure 2

##### Supplementary Figure 2

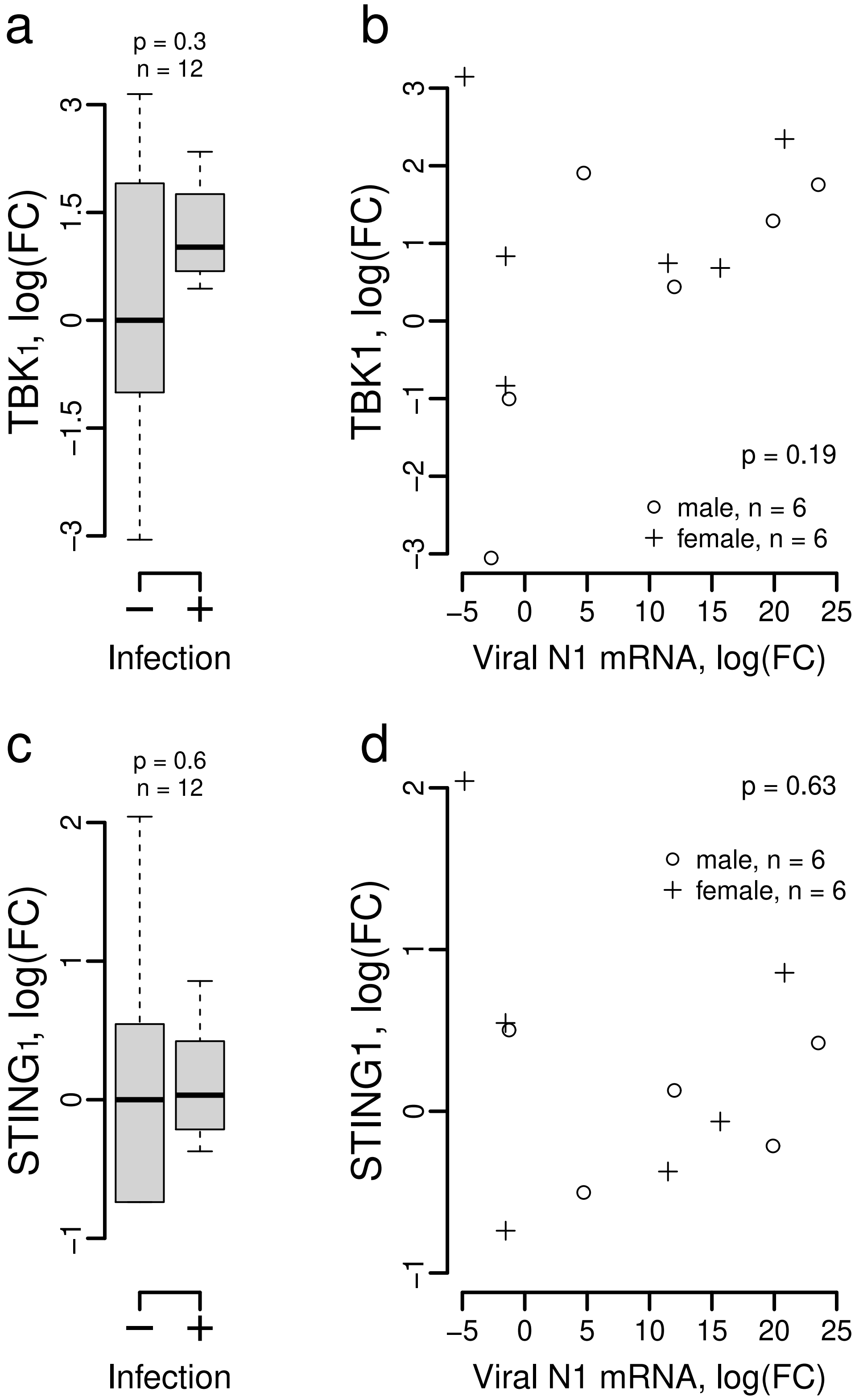
